# Cohort profile: The International epidemiology Databases to Evaluate AIDS (IeDEA) in sub-Saharan Africa, 2012-2019

**DOI:** 10.1101/19010231

**Authors:** Frédérique Chammartin, Cam Ha Dao Ostinelli, Kathryn Anastos, Antoine Jaquet, Ellen Brazier, Steven Brown, François Dabis, Mary-Ann Davies, Stephany N Duda, Karen Malateste, Denis Nash, Kara K Wools-Kaloustian, Per M von Groote, Matthias Egger

## Abstract

**Purpose:** The objectives of the International epidemiology Databases to Evaluate AIDS (IeDEA) are to (i) evaluate the delivery of combination antiretroviral therapy (ART) in children, adolescents and adults in sub-Saharan Africa, (ii) to describe ART regimen effectiveness, durability and tolerability, (iii) to examine HIV-related comorbidities and co-infections, and (iv) to examine the pregnancy- and HIV-related outcomes of women on ART and their infants exposed to HIV or antiretroviral therapy in utero or via breastmilk.

**Participants:** IeDEA is organized in four regions (Central, East, Southern and West Africa), with 240 treatment and care sites, six data centres at African, European and US universities, and almost 1.4 million children, adolescents and adult people living with HIV (PLWHIV) enrolled.

**Findings to date:** The data include socio-demographic characteristics, clinical outcomes, opportunistic events, treatment regimens, clinic visits and laboratory measurements. They have been used to analyse outcomes in people living with HIV-1 or HIV-2 who initiate ART, including determinants of mortality, of switching to second-line and third-line ART, drug resistance, loss to follow-up and the immunological and virological response to different ART regimens. Programme-level estimates of mortality have been corrected for loss to follow-up. We examined the impact of co-infection with hepatitis B and C, and the epidemiology of different cancers and of (multi-drug resistant) tuberculosis, renal disease and of mental illness. The adoption of “Treat All”, making ART available to all PLWHIV regardless of CD4^+^ cell count or clinical stage was another important research topic.

**Future plans:** IeDEA has formulated several research priorities for the “Treat All” era in sub-Saharan Africa. It recently obtained funding to set up sentinel sites where additional data are prospectively collected on cardiometabolic risks factors as well as mental health and liver diseases, and is planning to create a drug resistance database.

## INTRODUCTION

The introduction of combination antiretroviral therapy (ART) in sub-Saharan Africa from 2004 onwards has substantially improved the prognosis of HIV-1 infection, with a decline in AIDS-related deaths [1] and a decline in the incidence of new HIV-1 infections [2]. However, in many settings HIV/AIDS is still a public health threat. An estimated 1.8 million new infections occurred in 2017 and almost a million adult and child deaths were due to HIV, most of them in sub-Saharan Africa [2].

The World Health Organization (WHO), the Joint United Nations Programme on HIV/AIDS (UNAIDS) and many of the countries most heavily affected by the HIV epidemic have committed to ending HIV/AIDS as a major public health problem by 2030.[1] Targets to be reached by 2020 include that 90% of people living with HIV (PLWHIV) be aware of their status, 90% of those diagnosed initiate ART, and 90% of those on ART achieve undetectable viral loads (the 90-90-90 targets) [3]. Progress towards these goals has been substantial, especially in the most affected regions of Eastern and Southern Africa. The latest estimates suggested that the two regions are on track and had achieved 76%, 79% and 83% of the 90-90-90 targets, respectively in 2017 [4]. West and Central Africa face difficulties with the first 90% target and only an estimated 42% of PLWHIV in these regions were aware of their status [4].

Established more than ten years ago by the National Institutes of Health (NIH), the International epidemiology Databases to Evaluate AIDS (IeDEA) are a global cohort collaboration that collects HIV/AIDS data from HIV care and treatment programs, including in sub-Saharan Africa. The regional IeDEA data centres consolidate, curate and analyse data to evaluate the outcomes of people living with HIV/AIDS and monitor progress. The first years of the cohorts in sub-Saharan Africa were described previously [5]; here we provide an update on methods, key data and future plans.

## COHORT DESCRIPTION

In 2006 the National Institute of Allergy and Infectious Diseases (NIAID) sought applications for a global consortium structured through regional centres to pool clinical and epidemiological data on PLWHIV, in order to address questions that could not be answered by individual cohorts [5]. IeDEA covers seven geographic regions, namely North America, the Caribbean and Central/South America, the Asia-Pacific and four regions in sub-Saharan Africa: West Africa, Central Africa, East Africa and Southern Africa. The project was initially funded for a 5-year period and has since been extended twice, with the current funding cycle ending in 2021.

### Settings and number of PLWHIV enrolled

To date, the African regions of IeDEA received data from 240 HIV care and treatment facilities in 19 sub-Saharan African countries (Figure 1A). Close to 1,400,000 PLWHIV who initiated ART in sub-Saharan Africa are included (Figure 1B), of whom over 680,000 are currently in care. In East and Southern Africa, both urban and rural facilities are well represented, while in Central and West Africa, urban facilities dominate. Facilities are predominately public (95%) and operated at the primary or secondary care level, with the exception of West Africa where 70% of facilities are at the tertiary level of care (Table 1).

**Table 1:** Characteristics of 240 facilities providing ART in the African regions if the IeDEA (source: site assessment survey 2017).

|  | West Africa | Central Africa | East Africa | Southern Africa | All regions (%) |
| --- | --- | --- | --- | --- | --- |
| **No of active facilities** | 17 | 19 | 72 | 132 | 240 |
| **Location** |  |  |  |  |  |
| Urban | 17 | 19 | 19 | 65 | 120 (50) |
| Rural | 0 | 0 | 52 | 67 | 119 (50) |
| **Level of care** |  |  |  |  |  |
| Primary | 4 | 12 | 49 | 107 | 172 (74) |
| Secondary | 1 | 0 | 16 | 20 | 37 (16) |
| Tertiary | 12 | 7 | 6 | 5 | 24 (10) |
| **Type of facility** |  |  |  |  |  |
| Public | 12 | 16 | 69 | 128 | 225 (95) |
| Private | 2 | 3 | 3 | 4 | 12 (5) |
| **Viral load** |  |  |  |  |  |
| Routine testing | 12 | 16 | 46 | 74 | 148 (62) |
| Results available within 15 days | 6 | 13 | 11 | 22 | 52 (22) |
| Results available within 16-30 days | 7 | 4 | 11 | 104 | 126 (53) |
| Tests performed onsite | 8 | 6 | 4 | 3 | 21 (9) |
| Tests performed offsite | 6 | 13 | 53 | 127 | 199 (83) |
| **CD4 monitoring** |  |  |  |  |  |
| Routine testing | 14 | 6 | 9 | 97 | 126 (53) |
| Results available same day | 10 | 8 | 21 | 17 | 56 (23) |
| Tests performed onsite | 11 | 8 | 23 | 51 | 93 (39) |
| Tests performed offsite | 3 | 11 | 35 | 71 | 120 (50) |
| **HIV-1 genotypic drug resistance** |  |  |  |  |  |
| Routine testing | 2 | 2 | 10 | 48 | 62 (26) |
| **Routine tracing of LTFU** |  |  |  |  |  |
| Yes | 13 | 19 | 56 | 93 | 181 (75) |
| No | 1 | 0 | 1 | 37 | 39 (16) |
| **Tracing method\*** |  |  |  |  |  |
| Phone | 14 | 18 | 57 | 110 | 199 |
| SMS/mail/email | 2 | 2 | 10 | 14 | 28 |
| Home visit | 9 | 17 | 52 | 129 | 207 |
| **Medication disruption/stock outs last 12 m** |  |  |  |  |  |
| Pharmacy available on site | 14 | 19 | 56 | 96 | 185 |
| First-line ART | 5 | 2 | 20 | 32 | 59 |
| Second-line ART | 5 | 6 | 18 | 16 | 45 |

\* Sites may use more than 1 method

**Figure 1:**
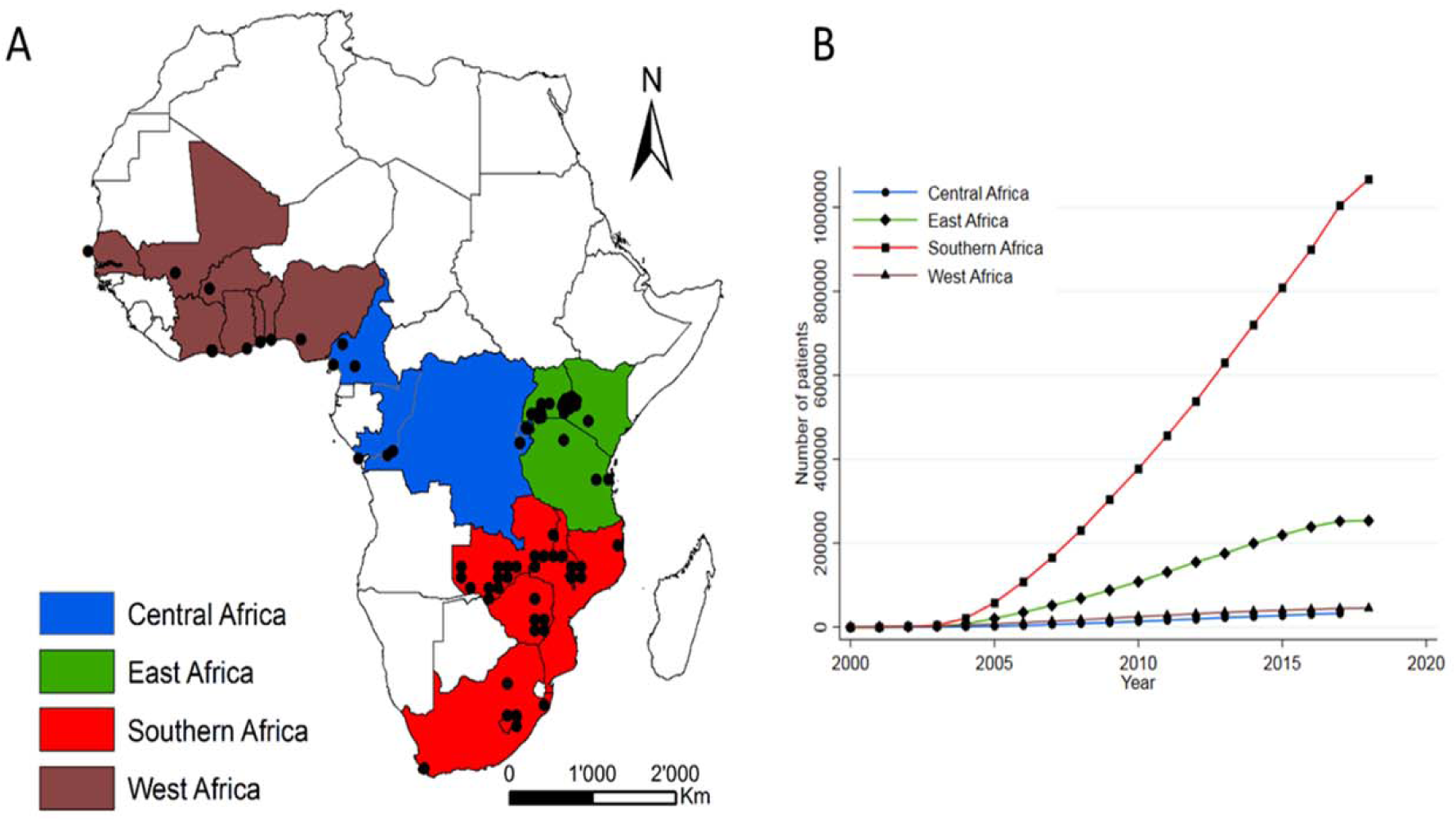
Map of the 240 active facilities participating in the four African regions of the International epidemiology Databases to Evaluate AIDS (A), together with cumulative numbers of patients starting antiretroviral therapy (B).

### Data collection at individual and site level

Since its inception, IeDEA has collected routine clinical data of PLWHIV followed under treatment, which includes socio-demographic characteristics, clinical outcomes, opportunistic events, treatment regimens, clinic visits and laboratory measurements. More recently, the IeDEA network has developed a Data Exchange Standard (DES) protocol (see www.iedeades.org) with 25 data tables, which include a total of 228 unique variables (36 compulsory and 192 additional variables) and within-region unique patient research identifiers. Standardized data collection is supported by eight codebooks, including the Anatomical Therapeutic Chemical (ATC) classification for drugs and lists with codes for reasons for stopping treatment, for dropping out of the cohort, for mode of HIV infection, country, type and site of comorbidities, laboratory measurements and units of measurements and type of viral load assay. The collection, management and sharing of data is facilitated by the Harmonist toolkit, a software and standards package that supports research projects through the Research Electronic Data Capture (REDCap) system [6].

In recent years, site assessments and site surveys have been conducted on a regular basis to collect up-to-date information related to available clinical service and care models in the participating facilities. For example, a study compared the characteristics and comprehensiveness of adult HIV care and treatment programmes in sub-Saharan Africa with programmes in the Americas and Asia-Pacific region [7]. Other studies examined the management of mental health and substance use disorders [8], or the diagnostic and screening practices for (drug resistant) tuberculosis in adult and paediatric patients [9–11]. Furthermore, the routine data collected by participating sites have been enriched in some countries by linking the routine HIV databases to cancer registries [12,13], vital registries [14] or administrative databases [15].

### Trends in CD4 cell count and viral load measurements

While WHO continues to recommend a CD4 cell count before starting ART to inform the management of advanced disease and differentiated care in the Treat All era, it also recommends that CD4 testing be replaced by viral load measurement for monitoring of treatment and identification of treatment failure [16]. Figure 2 shows that in Southern and West Africa, the number of CD4 measurements tended to be stable over time, despite an increasing number of PLWHIV in care, while in East and Central Africa the number of CD4 measurements dropped. At present, 53% of the active facilities reported routine CD4 testing and 65% routine viral load testing (Table 1). The United States President’s Emergency Plan for AIDS Relief (PEPFAR), which provides substantial funding for AIDS treatment, care and prevention in countries most affected by the epidemic, has progressively reduced its support for CD4 testing [17].

**Figure 2:**
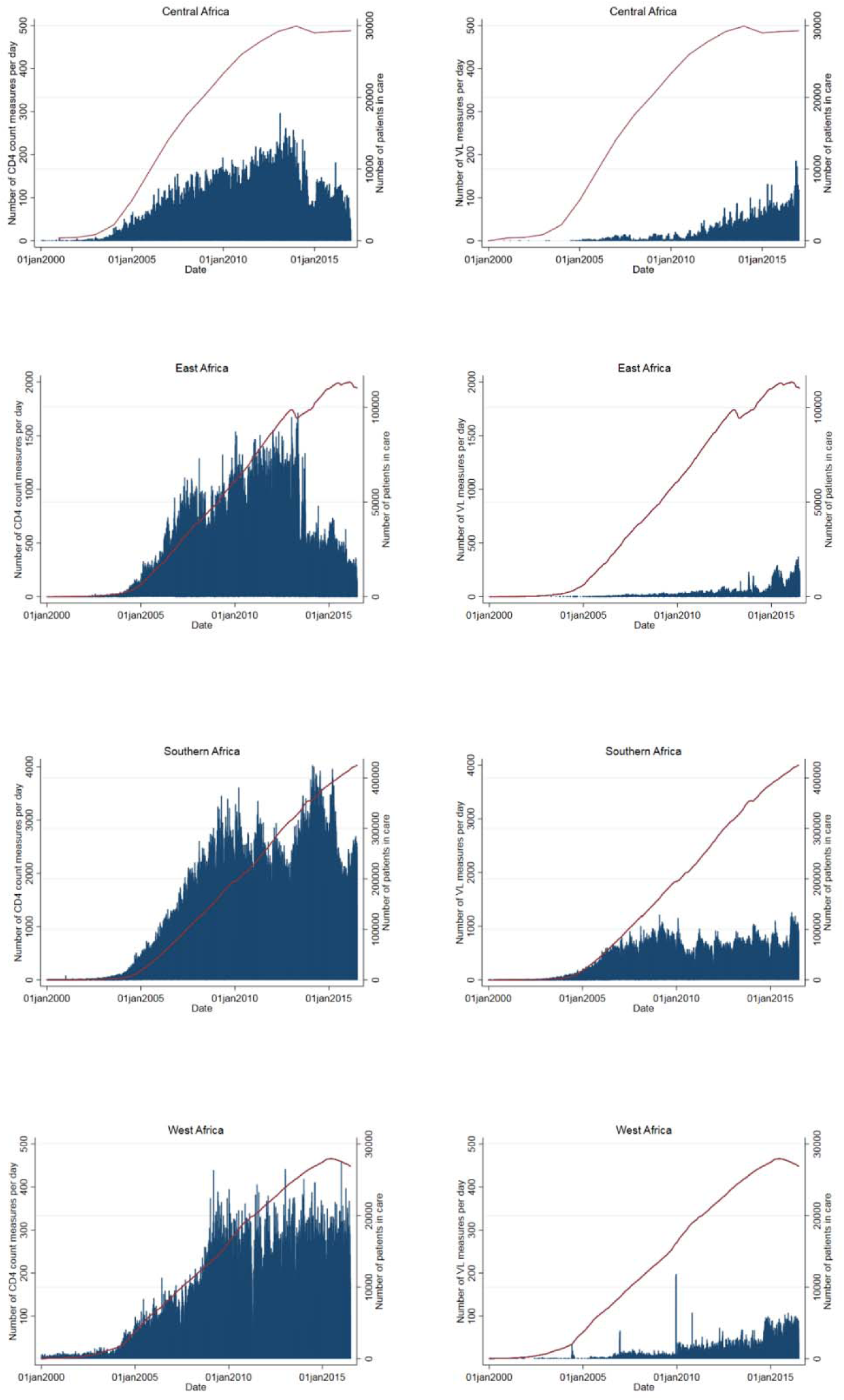
Daily number of CD4 cell counts (left) and viral load measurements (right) over time (bar chart) and the cumulative number of patients in care (red line).

### Trends in antiretroviral therapy

Until recently, the recommended first-line ART regimen in sub-Saharan Africa consisted of two nucleoside reverse transcriptase inhibitors and one non-nucleoside reverse transcriptase inhibitor (2NRTIs+1NNRTI). The combination of tenofovir (TDF), lamivudine (3TC) (or emtricitabine (FTC)) and efavirenz (EFV) is the current treatment of choice. The phasing out of stavudine (D4T) and nevirapine (NVP) was almost complete in 2014 (Table 2). East Africa and Southern Africa are currently rolling out dolutegravir (DTG), an integrase inhibitor with a high barrier to resistance [18][19]. Due to concerns about an increased risk of neural tube defects if taken during pregnancy[20], the roll-out to women has been delayed or limited in some settings. Of note, drug stock-outs in the last 12 months were reported by 59 facilities for first-line drugs, and by 45 for second-line drugs (Table 1).

**Table 2:**
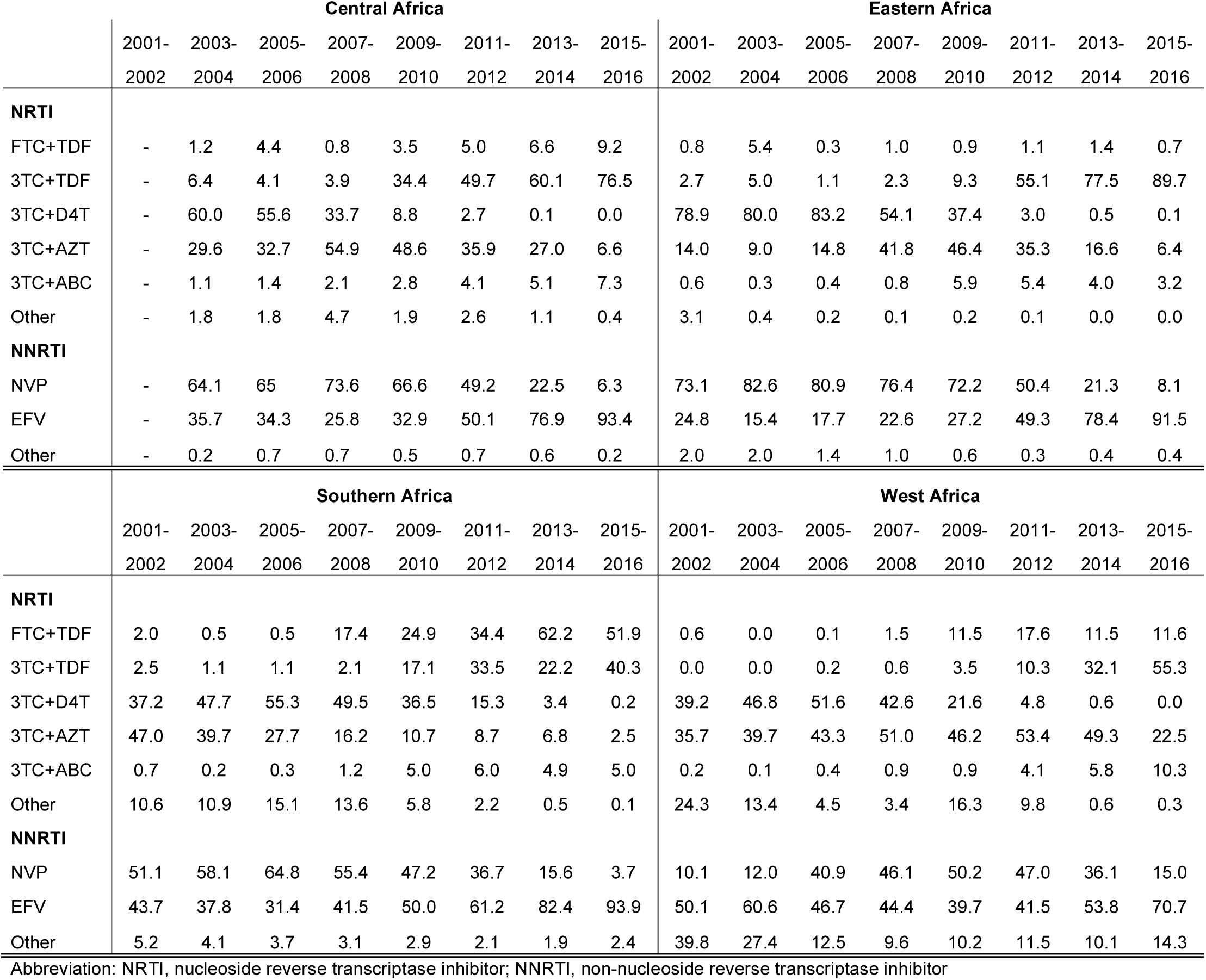
Proportion of patients with different nucleoside and non-nucleoside reverse transcriptase inhibitors regimen at ART start over time, by region.

|  | Central Africa |  |  |  |  |  |  |  | Eastern Africa |  |  |  |  |  |  |  |
| --- | --- | --- | --- | --- | --- | --- | --- | --- | --- | --- | --- | --- | --- | --- | --- | --- |
|  | 2001-2002 | 2003-2004 | 2005-2006 | 2007-2008 | 2009-2010 | 2011-2012 | 2013-2014 | 2015-2016 | 2001-2002 | 2003-2004 | 2005-2006 | 2007-2008 | 2009-2010 | 2011-2012 | 2013-2014 | 2015-2016 |
| **NRTI** |  |  |  |  |  |  |  |  |  |  |  |  |  |  |  |  |
| FTC+TDF | - | 1.2 | 4.4 | 0.8 | 3.5 | 5.0 | 6.6 | 9.2 | 0.8 | 5.4 | 0.3 | 1.0 | 0.9 | 1.1 | 1.4 | 0.7 |
| 3TC+TDF | - | 6.4 | 4.1 | 3.9 | 34.4 | 49.7 | 60.1 | 76.5 | 2.7 | 5.0 | 1.1 | 2.3 | 9.3 | 55.1 | 77.5 | 89.7 |
| 3TC+D4T | - | 60.0 | 55.6 | 33.7 | 8.8 | 2.7 | 0.1 | 0.0 | 78.9 | 80.0 | 83.2 | 54.1 | 37.4 | 3.0 | 0.5 | 0.1 |
| 3TC+AZT | - | 29.6 | 32.7 | 54.9 | 48.6 | 35.9 | 27.0 | 6.6 | 14.0 | 9.0 | 14.8 | 41.8 | 46.4 | 35.3 | 16.6 | 6.4 |
| 3TC+ABC | - | 1.1 | 1.4 | 2.1 | 2.8 | 4.1 | 5.1 | 7.3 | 0.6 | 0.3 | 0.4 | 0.8 | 5.9 | 5.4 | 4.0 | 3.2 |
| Other | - | 1.8 | 1.8 | 4.7 | 1.9 | 2.6 | 1.1 | 0.4 | 3.1 | 0.4 | 0.2 | 0.1 | 0.2 | 0.1 | 0.0 | 0.0 |
| **NNRTI** |  |  |  |  |  |  |  |  |  |  |  |  |  |  |  |  |
| NVP | - | 64.1 | 65 | 73.6 | 66.6 | 49.2 | 22.5 | 6.3 | 73.1 | 82.6 | 80.9 | 76.4 | 72.2 | 50.4 | 21.3 | 8.1 |
| EFV | - | 35.7 | 34.3 | 25.8 | 32.9 | 50.1 | 76.9 | 93.4 | 24.8 | 15.4 | 17.7 | 22.6 | 27.2 | 49.3 | 78.4 | 91.5 |
| Other | - | 0.2 | 0.7 | 0.7 | 0.5 | 0.7 | 0.6 | 0.2 | 2.0 | 2.0 | 1.4 | 1.0 | 0.6 | 0.3 | 0.4 | 0.4 |
|  | Southern Africa |  |  |  |  |  |  |  | West Africa |  |  |  |  |  |  |  |
|  | 2001-2002 | 2003-2004 | 2005-2006 | 2007-2008 | 2009-2010 | 2011-2012 | 2013-2014 | 2015-2016 | 2001-2002 | 2003-2004 | 2005-2006 | 2007-2008 | 2009-2010 | 2011-2012 | 2013-2014 | 2015-2016 |
| **NRTI** |  |  |  |  |  |  |  |  |  |  |  |  |  |  |  |  |
| FTC+TDF | 2.0 | 0.5 | 0.5 | 17.4 | 24.9 | 34.4 | 62.2 | 51.9 | 0.6 | 0.0 | 0.1 | 1.5 | 11.5 | 17.6 | 11.5 | 11.6 |
| 3TC+TDF | 2.5 | 1.1 | 1.1 | 2.1 | 17.1 | 33.5 | 22.2 | 40.3 | 0.0 | 0.0 | 0.2 | 0.6 | 3.5 | 10.3 | 32.1 | 55.3 |
| 3TC+D4T | 37.2 | 47.7 | 55.3 | 49.5 | 36.5 | 15.3 | 3.4 | 0.2 | 39.2 | 46.8 | 51.6 | 42.6 | 21.6 | 4.8 | 0.6 | 0.0 |
| 3TC+AZT | 47.0 | 39.7 | 27.7 | 16.2 | 10.7 | 8.7 | 6.8 | 2.5 | 35.7 | 39.7 | 43.3 | 51.0 | 46.2 | 53.4 | 49.3 | 22.5 |
| 3TC+ABC | 0.7 | 0.2 | 0.3 | 1.2 | 5.0 | 6.0 | 4.9 | 5.0 | 0.2 | 0.1 | 0.4 | 0.9 | 0.9 | 4.1 | 5.8 | 10.3 |
| Other | 10.6 | 10.9 | 15.1 | 13.6 | 5.8 | 2.2 | 0.5 | 0.1 | 24.3 | 13.4 | 4.5 | 3.4 | 16.3 | 9.8 | 0.6 | 0.3 |
| **NNRTI** |  |  |  |  |  |  |  |  |  |  |  |  |  |  |  |  |
| NVP | 51.1 | 58.1 | 64.8 | 55.4 | 47.2 | 36.7 | 15.6 | 3.7 | 10.1 | 12.0 | 40.9 | 46.1 | 50.2 | 47.0 | 36.1 | 15.0 |
| EFV | 43.7 | 37.8 | 31.4 | 41.5 | 50.0 | 61.2 | 82.4 | 93.9 | 50.1 | 60.6 | 46.7 | 44.4 | 39.7 | 41.5 | 53.8 | 70.7 |
| Other | 5.2 | 4.1 | 3.7 | 3.1 | 2.9 | 2.1 | 1.9 | 2.4 | 39.8 | 27.4 | 12.5 | 9.6 | 10.2 | 11.5 | 10.1 | 14.3 |

Abbreviation: NRTI, nucleoside reverse transcriptase inhibitor; NNRTI, non-nucleoside reverse transcriptase inhibitor

### Mortality and retention in care

In cohorts of PLWHIV who initiated ART in consecutive two-year periods from 2001 to 2016, mortality at 3 years declined substantially in all African IeDEA regions (Figure 3A). Loss to follow-up, defined as more than 90 days late to the next scheduled visit, remained substantial in all regions, and particularly high in Southern Africa (Figure 3B). Retention in care is key to the success of the public health approach to ART. Loss to follow-up has been an important issue for IeDEA, and activities to trace PLWHIV not returning to the clinic have increased in recent years. At present, tracing of PLWHIV on ART who were lost to follow-up is in place in 89% of the active facilities; 75% have implemented it routinely (Table 1). Tracing methods vary widely across facilities and include phone calls and home visits by clinic staff or community health workers.

**Figure 3.**
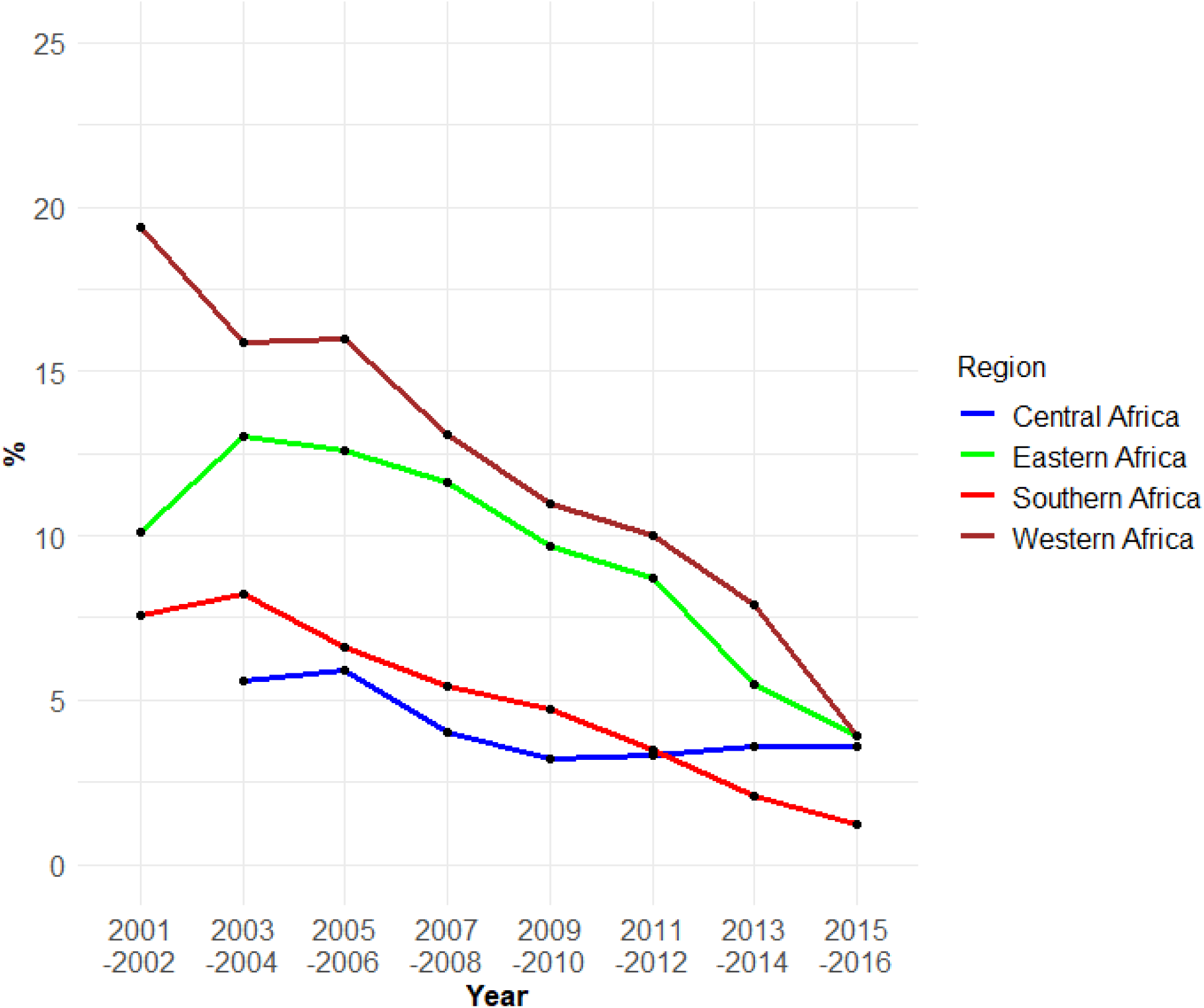
Trends in mortality (A) and loss to follow (B), 2001 to 2016.

### Patient and public involvement

IeDEA is based on the collection of routine clinical data and no patients were involved in developing the research question, outcome measures and overall design of the collaboration. Due to the anonymous nature of the data, we cannot disseminate the results of analyses of the data directly to study participants.

## KEY RESEARCH AREAS AND PUBLICATIONS

Over 500 publications in MEDLINE acknowledge funding from a core grant from the NIH to one or several African IeDEA regions, and these publications have been cited over 10,000 times. Multiregional projects are developed in IeDEA working groups, which currently address eight clinical areas: (i) cancer, (ii) ART outcomes, (iii) hepatitis, (iv) mental health, (v) mother-infant and paediatrics, (vi) renal disease, (vii) substance use and (viii) tuberculosis. Multiregional research concepts are discussed in the Executive Committee of IeDEA, revised and approved or rejected. In recent years, several analyses were done in collaboration with WHO or UNAIDS [21–23]. The number of publications reporting multi-regional analyses from several African IeDEA regions increased over time, from one such publication in 2007 to 24 multiregional publications in 2018. Some of the key studies are summarised below, with a focus on more recent and on multiregional analyses.

### Treatment outcomes in adults, adolescents, children and pregnant women

Several studies examined outcomes in people living with HIV-1 or HIV-2 who initiate ART, including determinants of mortality, of switching to second-line and third-line ART, drug resistance, loss to follow-up and the immunological and virological response to different ART regimens [24–37]. For example, the African IeDEA regions contributed importantly to a large-scale analysis of outcomes in adolescents living with perinatally acquired HIV, which showed that HIV-associated mortality during adolescence was substantially higher in sub-Saharan Africa, South and Southeast Asia, and South America and the Caribbean than in Europe [36]. A similar analysis of adolescents living with HIV showed that mortality and loss to follow-up were worse among those entering care at 15 years or older [37]. The authors concluded that adolescents must be evaluated separately from younger children and adults to identify population-specific reasons for death and loss to follow-up [37].

### Programme-level mortality

It became clear early on during the scale-up of ART in sub-Saharan Africa, that loss to follow-up of patients initiating ART was substantial [38], and that mortality among patients lost was higher than among patients remaining in care [39]. Ignoring loss to follow-up might thus bias programme-level estimates of mortality, and much effort has gone into correcting programme-level mortality for loss to follow-up [40–46]. For example, an analysis of all four African regions showed that when analysing the uncorrected data observed in the clinics, 52% of adults and children were retained on ART, 42% were lost to follow-up and 6% had died 5 years after ART initiation [46]. After accounting for undocumented deaths and self-transfers, an estimated 67% of patients were retained on ART, 19% had stopped ART and 15% had died [46].

### Co-infections and co-morbidities

IeDEA investigators have examined the prevalence and impact of co-infection with hepatitis B and C, and the epidemiology of different cancers and of (multi-drug resistant) tuberculosis, renal disease and of mental illness [8,47–58]. For example, an analysis of IeDEA data and data from the Collaboration of Observational HIV Epidemiological Research in Europe (COHERE) [59] showed that children living with HIV from sub-Saharan Africa, but not those from Europe or Asia had a high risk of developing Kaposi sarcoma after starting ART [58]. Similarly, a recent analysis of IeDEA and COHERE data showed that compared to European women, rates of cervical cancer were 11 times higher in South Africa [50]. A recent multiregional study of multi-drug resistant tuberculosis included HIV positive and HIV negative adults with tuberculosis from seven high-burden countries (Côte d’Ivoire, Democratic Republic of the Congo, Kenya, Nigeria, South Africa, Peru, and Thailand). Molecular or phenotypic drug susceptibility testing was done locally and at a reference laboratory. The results showed that inaccurate local drug susceptibility testing led to under-treatment of drug-resistant tuberculosis and increased mortality [54].

### The challenge of “Treat All”

Nearly all countries in sub-Saharan Africa have now adopted national polices to offer ART to all PLWHIV regardless of CD4 cell count or clinical stage (‘Treat All’), in order to meet the UNAIDS 90-90-90 targets. In 2011, Malawi was one of the first countries to implement such a strategy for the prevention of mother-to-child transmission (PMTCT), recommending ART for pregnant and breastfeeding women living with HIV, regardless of CD4 cell count or WHO clinical stage (“Option B+”). An IeDEA analysis of the Malawian experience showed that poor retention in care was a problem in many facilities, with early loss to follow-up particularly high in facilities with a high patient volume and in patients who start ART during pregnancy on the day of HIV diagnosis [35]. More recently, IeDEA investigators used regression discontinuity analysis to examine changes in rapid HIV treatment initiation after national *“*Treat All*”* policy adoption in six countries (Burundi, Kenya, Malawi, Rwanda, Uganda, Zambia) [60]. They showed a strong and sustained effect of the adoption of “Treat All” policies on ART initiation within 30 days of enrollment in HIV care in all six countries [60].

## THE FUTURE

At the end of 2018, IeDEA published a consensus statement [61] and a journal supplement [62] on research priorities to inform the implementation of the “Treat All” policy in children and adolescents[63], pregnant and post-partum women [64], and for mental health, substance use [65] and drug resistance [66]. These documents will guide IeDEA’s future research agenda in sub-Saharan Africa. Furthermore, the creation of an IeDEA Sentinel Research Network (IeDEA-SRN) will facilitate the collection of detailed data in selected IeDEA sites on cardio-metabolic risk factors (e.g. hypertension, diabetes, dyslipidaemia) liver disease (liver fibrosis and steatosis), mental health and substance use. In the East and Southern African regions, pharmacovigilance in pregnancy is being developed to assess the impact of ART on birth outcomes. A project involving all four African regions studies the cascade of screening for cervical cancer, while the establishment of the South African HIV Cancer Match (SAM) study of over ten million PLWHIV, from linkage of national laboratory with cancer registry data, will allow the study of less common cancers. Finally, the creation of a drug resistance database as a central repository for resistance tests performed in routine clinical care is another planned addition.

## COLLABORATIONS

The African regions of IeDEA have collaborated and continue to encourage collaborations with other consortia, cohort collaborations, the HIV modelling community and public health agencies as well as individuals wishing to use IeDEA data. Examples include work with COHERE [50,58,67,68], the Collaborative Initiative for Paediatric HIV Education and Research (CIPHER) collaboration [36,69] or the Measurement & Surveillance of HIV Epidemics (MeSH) consortium [22] as well as UNAIDS [22,23] and WHO [21]. Furher collaborations are welcome. Investigators wishing to work with the IeDEA data should contact the teams at the regional data centres (see www.iedea.org for contact details) and send a concept sheet for the analyses they are interested in performing and the variables that would be required. Anyone wishing to work with IeDEA must sign a data-use agreement.

## Data Availability

Investigators wishing to work with IeDEA data should contact the regional data centres (see www.iedea.org) and send a concept sheet for the analyses they are interested in performing and the variables that would be required. See www.iedeades.org for list of variables. Those wishing to work with the data must sign a data-use agreement.

## FOOTNOTES

### Acknowledgements

The authors wish to thank the clinical and administrative staff at the participating clinics. We are grateful to all PLWHIV who contributed to the IeDEA database.

### Contributors

FC, PMG, and ME conceptualised the study. FC and CHDO performed statistical analyses. FC and ME wrote the first draft of the paper. CHDO, KA, AJ, EB, SB, FD, MAD, SND, KM, BSM, DN, KWK, CY, PMG and ME contributed to interpreting the data and to the writing and revising of the manuscript.

### Funding

The International Epidemiology Databases to Evaluate AIDS (IeDEA) is supported by the U.S. National Institutes of Health’s National Institute of Allergy and Infectious Diseases, the Eunice Kennedy Shriver National Institute of Child Health and Human Development, the National Cancer Institute, the National Institute of Mental Health, the National Institute on Drug Abuse and Alcoholism, the National Institute of Diabetes and Digestive and Kidney Diseases, the Fogarty International Center, the National Library of Medicine, and the Office of the Director: Central Africa, U01AI096299; East Africa, U01AI069911; Southern Africa, U01AI069924; West Africa, U01AI069919. Informatics resources are supported by the Harmonist project, R24AI124872. ME was supported by special project funding (Grant No. 174281) from the Swiss National Science Foundation.

### Disclaimer

The contents are the responsibility of the authors and do not necessarily reflect the views of NIAD or the US Government. The funders had no role in study design, data collection and analysis, decision to publish or preparation of the manuscript.

### Competing interests

None declared.

### Ethics approval

Ethics Committee of the Canton of Bern, Switzerland.

### Provenance and peer review

Not commissioned; externally peer reviewed.

### Open Access

This is an Open Access article distributed in accordance with the Creative Commons Attribution Non Commercial (CC BY-NC 4.0) license, which permits others to distribute, remix, adapt, build upon this work non-commercially, and license their derivative works on different terms, provided the original work is properly cited and the use is non-commercial. See: http://creativecommons.org/licenses/by-nc/4.0/.

**Box: Strengths and limitations of this study**

- An important strength of the IeDEA cohort collaboration in sub-Saharan Africa is its large size, which allows analyses of outcomes of antiretroviral therapy (ART) in children, adolescents and pregnant and postpartum women, across diverse settings.
- The data reflect routine care across a wide range of real-world settings during the scale up of ART in sub-Saharan Africa and thus provide a valuable platform to conduct operational and clinical research and to study temporal trends and the impact of changes in guidelines and other interventions.
- The development of a standardised Data Exchange Standard protocol has contributed to increase data quality, and data have been enriched by linkage to cancer registries, vital registries and administrative databases.
- Collaborations with the World Health Organization, UNAIDS, the mathematical modelling community and other consortia have ensured that the analyses of the African IeDEA regions contributed substantially to global health policy and decision making.
- Weaknesses include the limitations inherent in secondary use of routine clinical care data, with missing data, the lack of standardised follow-up visits, and substantial loss to follow-up resulting in unknown outcome.
- Additional challenges include the lack of population-based samples, including HIV negative controls or ART-naive comparison groups, and the lack of measurements such as non-communicable disease risk factors and outcomes. Dealing with risk factors that affect clinical decision making, and the need to control for (time-dependent) confounding can also be challenging.

## Notes

### Competing Interest Statement

The authors have declared no competing interest.

### Author Declarations

All relevant ethical guidelines have been followed and any necessary IRB and/or ethics committee approvals have been obtained.

Any clinical trials involved have been registered with an ICMJE-approved registry such as ClinicalTrials.gov and the trial ID is included in the manuscript.

